# DETERMINANTS AND CONSEQUENCES OF PATIENT AND HEALTH SYSTEM DELAYS IN TUBERCULOSIS DIAGNOSIS AND TREATMENT AMONG INDIVIDUALS AGED ≥15 YEARS AT KENYATTA NATIONAL HOSPITAL, NAIROBI, KENYA

**DOI:** 10.64898/2026.03.02.26347417

**Authors:** Magoba Ronald Arnold, Dominic Mogere, Dennis Magu, Mark Mudenyo

**Affiliations:** Mount Kenya University; Jomo Kenyatta University of Agriculture and Technology; Kenyatta University

**Keywords:** Tuberculosis, Patient delay, Health system delay, Diagnostic delay, Care-seeking pathways, Kenya

## Abstract

**Background:** Tuberculosis (TB) remains a leading cause of morbidity and mortality globally, particularly in low- and middle-income countries. Delays in diagnosis and treatment initiation significantly contribute to ongoing transmission, increased disease severity, and poor treatment outcomes. This study assessed the determinants and consequences of patient and health system delays among individuals aged 15 years and above at Kenyatta National Hospital, Nairobi, Kenya.

**Methods:** A retrospective cohort study was conducted among 127 TB patients aged ≥15 years. Data were collected using structured questionnaires and clinical record reviews to reconstruct timelines from symptom onset to treatment initiation. Patient delay was defined as the time from symptom onset to first healthcare contact, while health system delay referred to the time from first contact to confirmed diagnosis. Descriptive statistics, chi-square tests, and multivariable logistic regression were used to identify factors associated with delay. Results were reported as adjusted odds ratios (AORs) with 95% confidence intervals.

**Results:** The majority of participants were aged 40–49 years (32.3%), indicating a high burden among economically productive populations. Over half of the participants (59.8%) experienced prolonged patient delay exceeding two months. Key barriers to timely care-seeking included stigma (96.9%), fear of diagnosis (66.9%), and long distances to health facilities (31.5% residing >10 km away). Many participants initially sought care from informal providers or engaged in self-medication, contributing to complex and delayed care pathways. Bivariate analysis showed significant associations between delay and age (p = 0.032), distance to health facility (p = 0.015), and education level (p = 0.041). However, no independent predictors remained statistically significant after multivariable adjustment.

**Conclusion:** Patient and health system delays remain substantial challenges in TB control, driven by a combination of behavioral, social, and structural factors. Interventions should prioritize community awareness, stigma reduction, improved access to healthcare services, and strengthening of referral systems. Addressing these delays is critical to reducing transmission and improving TB outcomes in high-burden settings.

## INTRODUCTION

Tuberculosis (TB) remains a major global public health challenge and continues to be one of the leading causes of morbidity and mortality worldwide, particularly in low- and middle-income countries. According to the World Health Organization, an estimated 10.6 million people developed TB in 2022, with a significant burden concentrated in sub-Saharan Africa (WHO, 2023). Despite advances in diagnostic technologies and treatment strategies, TB control efforts remain constrained by delays in diagnosis and treatment initiation, which contribute to sustained transmission, increased disease severity, and poor treatment outcomes (WHO, 2023; Sreeramareddy et al., 2021).

Timely diagnosis and treatment are critical components of effective TB control. Delays in the care continuum are broadly categorized into patient delay and health system delay. Patient delay refers to the time between the onset of symptoms and the first contact with a healthcare provider, while health system delay refers to the time from initial contact to confirmed diagnosis and treatment initiation (Getnet et al., 2017; Sreeramareddy et al., 2021). Prolonged delays increase the duration of infectiousness, thereby facilitating ongoing community transmission and worsening clinical outcomes, including advanced disease presentation and increased mortality (WHO, 2023).

Multiple interrelated factors contribute to delays in TB diagnosis and treatment. At the individual level, limited awareness of TB symptoms, misinterpretation of early signs, and financial constraints often lead to delayed healthcare seeking (Getnet et al., 2017; Datiko et al., 2020). Social determinants such as stigma and fear of discrimination remain significant barriers, influencing care-seeking behavior and leading to concealment of symptoms. TB-related stigma continues to be widely documented as a critical obstacle to early diagnosis and treatment (Craig et al., 2017; Courtwright et al., 2019).

At the health system level, structural and operational challenges also contribute to delays. These include limited accessibility of healthcare services, long distances to facilities, inadequate diagnostic capacity, and inefficiencies in referral systems (WHO, 2023; Datiko et al., 2020). In many high-burden settings, healthcare systems are fragmented, requiring patients to navigate between public, private, and informal providers. This fragmentation often results in complex and non-linear care-seeking pathways, which further delay diagnosis and treatment (Sreeramareddy et al., 2021).

Care-seeking behavior in TB is frequently characterized by multiple steps before reaching appropriate diagnostic services. Patients often initiate care through self-medication, pharmacies, or traditional healers before presenting to formal healthcare facilities. These pathways reflect both patient preferences and systemic gaps in healthcare delivery. Each additional step introduces delays, increasing the risk of disease progression and continued transmission within communities (Datiko et al., 2020; Sreeramareddy et al., 2021).

Tuberculosis disproportionately affects individuals in economically productive age groups, amplifying both health and socioeconomic consequences. Adults within this demographic are more likely to engage in activities that increase exposure and transmission risk, while also experiencing significant economic impacts due to illness-related productivity losses (WHO, 2023; Furin et al., 2019). This underscores the dual burden of TB as both a biomedical and socioeconomic disease.

From a public health perspective, delays in TB diagnosis and treatment are best understood within the broader framework of social determinants of health. The Ottawa Charter for Health Promotion highlights the importance of creating supportive environments, strengthening community action, and improving access to essential health services. Addressing TB delays therefore requires a comprehensive approach that integrates community-level interventions, health system strengthening, and policy-level action, particularly in high-burden settings (WHO, 2023).

Although several studies have examined TB delays globally, there remains a need for context-specific evidence that captures the interaction between patient behavior, social determinants, and health system constraints in Kenya. Understanding these dynamics is essential for designing targeted, evidence-based interventions that improve early diagnosis and treatment outcomes.

This study therefore aimed to assess the determinants and consequences of patient and health system delays in TB diagnosis and treatment among individuals aged 15 years and above at Kenyatta National Hospital in Nairobi, Kenya. By examining care-seeking behavior, barriers to timely diagnosis, and pathways to care, the study seeks to generate evidence to inform strategies for reducing delays and strengthening TB control efforts in high-burden settings.

## METHODOLOGY

### Study Design

This study employed a **retrospective cohort design** to assess patient delay, barriers to care, and care-seeking pathways among individuals diagnosed with tuberculosis. The study utilized routinely collected clinical records, complemented by patient-reported information, to reconstruct timelines from symptom onset to diagnosis and initiation of treatment.

A retrospective cohort approach was appropriate for examining temporal relationships between exposures and outcomes using existing data. In this study, exposures included patient-related and health system factors associated with delays, while outcomes focused on time to diagnosis and treatment initiation. This design enabled the reconstruction of patient care pathways and identification of key points at which delays occurred. Retrospective cohort studies are widely applied in investigations of diagnostic and treatment delays due to their ability to provide practical and reliable epidemiological insights without the need for prolonged follow-up (1,2).

### Study Setting

The study was conducted in a high tuberculosis burden setting in Kenya, where TB remains a significant public health concern. The healthcare system is characterized by a mix of public facilities, private providers, pharmacies, and informal care options, all of which influence patient care-seeking behavior (1).

The study site was Kenyatta National Hospital, the largest national referral and teaching hospital in the country. The facility serves as a major center for TB diagnosis and management and receives referrals from across Kenya, including patients with advanced or complicated disease. As a tertiary institution with established TB programs and comprehensive patient records, the hospital provides an appropriate setting for examining diagnostic and treatment delays in a real-world context. Similar referral hospitals in high-burden settings are commonly used in delay studies due to their capacity to capture diverse patient populations and complex care pathways (2).

### Study Population

The study population comprised individuals aged 15 years and above with a confirmed diagnosis of pulmonary tuberculosis and receiving treatment at Kenyatta National Hospital. Participants were selected based on predefined eligibility criteria, including confirmed diagnosis and willingness to participate in the study.

The inclusion of individuals aged 15 years and above was considered appropriate, as this group represents adolescents and adults who exhibit distinct healthcare-seeking behaviors compared to children. Additionally, individuals within this age group contribute significantly to TB transmission due to higher levels of social interaction and mobility, making them a critical population for TB control interventions.

### Sample Size and Sampling Technique

A representative sample of TB patients was selected using systematic random sampling. The sample size was determined based on prevalence estimates from previous TB studies and adjusted for expected non-response rates to ensure statistical validity (Storla et al., 2008). Sample size calculation followed the Fisher et al. formula; a standard method used in health research for estimating proportions in populations.

The formula:

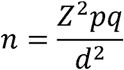

Where:

- *z* = 1.96(for 95% confidence level)
- *p* = 0.5(assumed proportion for maximum variability)
- *q* = 1 - *p*
- *d* = 0.05(margin of error)

This method is widely recommended because using **p = 0.5** ensures the largest possible sample size, making the study more statistically robust when the true proportion is unknown (Lwanga & Lemeshow, 1991).

A finite population correction was applied because the target population was less than 10,000, reducing the required sample size to **96**. However, the final sample was **128 participants**, which increases:

- Statistical power
- Precision of estimates
- Reliability of findings Sampling Procedure

A systematic sampling technique was used, which is appropriate in clinical settings where patients are listed in chronological or sequential order.

The process involved:

Identifying the sampling frame (TB register)
Calculating the sampling interval (K)
Selecting a random starting point
Selecting every Kth patient thereafter

Systematic sampling reduces selection bias while ensuring that the sample is spread evenly across the population (Kothari, 2004).

In addition, purposive sampling was used to select key informants. This is a qualitative sampling method where participants are intentionally chosen because they possess specific knowledge or experience relevant to the study. This approach is widely used in health systems research to capture in-depth contextual insights (Patton, 2015).

### Eligibility Criteria

Participants were eligible for inclusion if they were aged 15 years or older, had a confirmed diagnosis of pulmonary tuberculosis, and were actively receiving treatment at the study site during the study period. All participants provided informed consent prior to participation.

Participants were excluded if they had extra-pulmonary tuberculosis, as the diagnostic pathways and disease progression differ from pulmonary TB. Individuals with a history of previous TB treatment were also excluded to minimize potential bias related to recurrent or drug-resistant disease. In addition, patients who were too ill to participate at the time of data collection were excluded to ensure reliability of responses. Non-residents were also excluded to maintain consistency with the study context and ensure that findings reflected the local setting.

These eligibility criteria were applied to ensure a relatively homogeneous study population and to improve the internal validity of the findings (3).

### Data Collection

Data were collected using a structured questionnaire administered through face-to-face interviews. The tool captured information on socio-demographic characteristics, patient delay (defined as the time from symptom onset to first healthcare contact), barriers to care, and care-seeking pathways prior to diagnosis.

The questionnaire was adapted from previously validated tools used in tuberculosis research and was pre-tested prior to data collection to ensure clarity, consistency, and reliability. Minor adjustments were made following pre-testing to improve comprehension and flow (5).

### Quantitative Component

Quantitative data were obtained through a combination of retrospective chart review and structured questionnaires. Clinical records were reviewed to extract information on diagnosis timelines, treatment initiation dates, and relevant clinical characteristics. The structured questionnaires provided complementary data on socio-demographic factors, healthcare-seeking behavior, and knowledge related to tuberculosis. Participant recruitment and data collection were conducted over a seven-week period, commencing on 10/07/2022 and concluding on 30/08/2022, among individuals aged 15 years and above attending Kenyatta National Hospital for tuberculosis diagnostic and treatment services.

The use of multiple data sources allowed for triangulation of information, thereby enhancing the completeness and validity of the dataset (6).

### Bias and Limitations

Recall bias may have occurred, as the estimation of delay relied partly on patient self-report of symptom onset. To minimize this, interviews were conducted as close to the time of diagnosis as possible, and clinical records were used to validate reported timelines where available.

### Qualitative Component

A qualitative component was incorporated through Key Informant Interviews (KIIs) to provide deeper insight into patient experiences, health system barriers, and decision-making processes related to care-seeking. This approach complemented the quantitative findings by exploring underlying reasons for delays and contextual factors influencing healthcare utilization. Qualitative methods are particularly valuable in delay studies, as they help explain patterns observed in quantitative data (7).

### Measurement of Delays

Delays were defined and categorized in accordance with established global frameworks. Patient delay was defined as the time from onset of symptoms to first contact with a healthcare provider, while health system delay referred to the time from first contact to confirmed diagnosis. Total delay was calculated as the sum of these intervals. A threshold of more than 14 days was used to define prolonged delay, consistent with standard definitions in tuberculosis research (1,2).

### Data Analysis

Data were analyzed using SPSS version 25. Descriptive statistics were used to summarize participant characteristics. Associations between independent variables and delay outcomes were initially assessed using chi-square tests. Variables of interest were then included in binary logistic regression models to identify predictors of delay while controlling for potential confounders. Results were presented as adjusted odds ratios with corresponding 95% confidence intervals, and statistical significance was set at p < 0.05 (8).

Qualitative data were analyzed using thematic analysis. This involved systematic coding of transcripts, grouping of codes into categories, and development of themes that captured recurring patterns in participant responses. This approach allowed for structured interpretation of qualitative data and facilitated integration with quantitative findings (9).

### Reliability and Validity

Measures were taken to ensure the reliability and validity of the study. The data collection tools were pre-tested prior to use to enhance clarity and consistency. Standardized procedures were followed during data collection to minimize variability. These steps helped improve the accuracy and credibility of the findings (3).

### Ethical Considerations

The study was conducted in accordance with established ethical principles for human subject’s research. Participation was voluntary, and informed consent was obtained from all participants. Confidentiality of participant information was maintained throughout the study.

Ethical approval was obtained from relevant institutional review bodies, including the University of Nairobi/Kenyatta National Hospital and Mount Kenya University, as well as the National Commission for Science, Technology and Innovation. Adherence to these ethical standards was essential to ensure the protection of participants and the integrity of the research process (10).

## RESULTS

A total of 127 participants were included in the study. Females accounted for 51.2% (n = 65), while males comprised 48.8% (n = 62). The majority of participants were aged 40–49 years (32.3%), followed by 30–39 years (26.8%). Most participants had attained primary (31.5%) or secondary education (27.6%), and 43.3% were self-employed.

More than half of the participants (59.8%) reported seeking care after more than two months of symptom onset. Only 17.3% sought care within the first month.

A high proportion of participants reported perceived stigma (96.9%), while 66.9% reported fear following diagnosis. Regarding access, 31.5% of participants resided more than 10 km from a health facility.

Participants reported multiple initial points of care before diagnosis. These included self-medication, pharmacies, and informal providers prior to presentation at formal healthcare facilities.

Bivariate analysis showed that age group (p = 0.032), distance to health facility (p = 0.015), and education level (p = 0.041) were significantly associated with delay. Sex was not significantly associated (p = 0.210).

In multivariable logistic regression analysis, none of the variables remained statistically significant predictors of delay after adjustment (p > 0.05).

### Age Distribution of Participants

The age distribution of participants revealed that the majority were within the 40–49-year age group. This indicates that TB disproportionately affects individuals in economically productive age groups, consistent with global TB epidemiology (World Health Organization, 2023; Furin et al., 2019). This finding aligns with global epidemiological evidence demonstrating that TB disproportionately affects adults in working age groups due to increased social exposure, occupational risks, and cumulative lifetime exposure to *Mycobacterium tuberculosis* (World Health Organization, 2023; Furin et al., 2019). Individuals in this age bracket often engage in higher levels of mobility and social interaction, increasing the probability of both infection and transmission. Furthermore, TB in this age group has broader socioeconomic implications, including loss of productivity, increased dependency, and financial strain on households. This reinforces the need for targeted TB interventions focused on economically active populations to interrupt transmission chains.

Age distribution was analyzed descriptively using frequencies and percentages. No inferential test is required for this variable as it describes the study population. The predominance of TB among individuals aged 40–49 years aligns with global TB epidemiology, where disease burden is concentrated among economically productive adults (World Health Organization, 2023; Furin et al., 2019). This age group is more likely to experience repeated exposure, occupational mobility, and social contact, increasing infection risk. This finding reinforces the epidemiological concept that TB is both a **biological and socioeconomic disease**, disproportionately affecting individuals in their most productive years.

### Patient Delay Distribution

The study found that a substantial proportion of participants experienced prolonged patient delay, with many seeking care more than two months after symptom onset. This indicates significant delays in healthcare seeking, which may contribute to ongoing disease transmission and worsened clinical outcomes (Storla et al., 2008). Patient delay remains a critical barrier in TB control and is strongly associated with ongoing community transmission and poor clinical outcomes. Delays of this magnitude are consistent with findings from systematic reviews indicating that diagnostic delays are common in low- and middle-income countries (Storla et al., 2008; Sreeramareddy et al., 2014).

Several factors contribute to this delay. First, limited awareness of TB symptoms leads individuals to normalize chronic cough and delay seeking care. Second, economic constraints often discourage early healthcare seeking due to anticipated costs. Third, reliance on self-medication and informal care providers further prolongs the diagnostic timeline.

Prolonged patient delay increases bacterial load and infectiousness, thereby sustaining TB transmission within communities and complicating disease control efforts. Prolonged patient delay is a well-documented challenge in TB control and contributes significantly to disease transmission and poor outcomes (Storla et al., 2008; Sreeramareddy et al., 2014).

Delays are often influenced by:

- Limited TB symptom awareness
- Perceived mildness of symptoms
- Economic barriers
- Reliance on self-medication

The persistence of delayed care-seeking suggests a need for **community-level interventions**, as patient behavior plays a central role in TB control.

### Barriers to Care

The results showed that stigma was the most commonly reported barrier to care. Fear of diagnosis and social consequences also significantly influenced care-seeking behavior. In addition, geographical barriers such as distance to healthcare facilities were reported, highlighting structural challenges in accessing TB services (Courtwright & Turner, 2010; Lönnroth et al., 2009). Stigma remains a well-documented barrier to TB diagnosis and treatment. It operates at both social and psychological levels, influencing patients’ perceptions and behaviors. Individuals may avoid seeking care due to fear of discrimination, social exclusion, or loss of employment (Courtwright & Turner, 2010; Craig et al., 2017).

Fear of diagnosis is often intertwined with stigma, as TB is historically associated with poverty and social marginalization. This leads to concealment of symptoms and delayed engagement with healthcare services. Geographical barriers, including long distances to healthcare facilities and limited transportation options, further exacerbate access challenges, particularly in resource-limited settings (Lönnroth et al., 2009). These structural barriers highlight inequities in healthcare access and underscore the importance of decentralizing TB services. Collectively, these barriers demonstrate that TB is not only a biomedical disease but also a social condition influenced by structural determinants of health.

The most commonly reported barriers included:

- **Stigma**
- **Fear of diagnosis**
- **Geographical distance to health facilities**

### Statistical Analysis

- Barriers were analyzed using **frequency distributions**
- Association between barriers and delay was tested using:

- Chi-square test (χ²)
- Logistic regression for predictors of delay

Stigma remains a significant barrier to TB care and is strongly associated with delayed diagnosis and treatment (Courtwright & Turner, 2010; Craig et al., 2017).

Fear of social exclusion and discrimination discourages individuals from seeking care early, leading to concealment of symptoms. Additionally, geographical barriers limit access to diagnostic and treatment services, particularly in resource-constrained settings (Lönnroth et al., 2009). These findings highlight that TB control is not solely dependent on clinical services but also on **social determinants of health and health system accessibility**.

### Care-Seeking Pathways

The care-seeking pathways demonstrated that many participants initially engaged in self-medication before seeking formal healthcare services. Others first visited informal providers, including pharmacies and traditional healers, before eventually accessing formal healthcare facilities. This multi-step pathway contributed to delays in diagnosis and treatment initiation (Hoa et al., 2013). This fragmented care-seeking pattern is consistent with studies in similar settings, where patients initially consult pharmacies, traditional healers, or engage in self-treatment before accessing formal health systems (Hoa et al., 2013; Sreeramareddy et al., 2014).

Each step in this pathway introduces delays in diagnosis and treatment initiation. Informal providers often lack the capacity to diagnose TB or refer patients appropriately, resulting in missed opportunities for early detection. This multi-step pathway reflects both patient-level behavior and health system inefficiencies. It highlights gaps in healthcare integration, weak referral systems, and limited community-level TB screening.

Strengthening the role of community health systems and integrating informal providers into TB control strategies could significantly reduce diagnostic delays and improve outcomes. Participants followed **multi-step care-seeking pathways**, often starting with:

- Self-medication
- Pharmacies
- Traditional healers
- Later transitioning to formal healthcare facilities

Care-seeking pathways in TB are often complex and non-linear, reflecting both patient behavior and health system fragmentation (Hoa et al., 2013). Informal providers often act as the first point of contact but lack TB diagnostic capacity, leading to missed opportunities for early detection. This contributes to diagnostic delays and sustained transmission within communities. Improving integration between informal and formal healthcare providers is essential for early case detection and reducing delays.

## INTEGRATED SCIENTIFIC SYNTHESIS (HIGH-LEVEL DISCUSSION)

Across all variables, the findings demonstrate that TB delays are driven by an interaction of:

- **Individual factors** (age, knowledge, behavior)
- **Social factors** (stigma, fear)
- **System factors** (accessibility, referral gaps)

This aligns with TB control frameworks emphasizing that delays occur at both:

- Patient level
- **Health system level** (Lönnroth et al., 2009)

The persistence of these delays suggests that TB control strategies must adopt a **multi-sectoral approach**, combining:

- Community awareness
- Health system strengthening
- Social protection measures

**Table 1** presents the socio-demographic characteristics of the participants. Females constituted a slightly higher proportion (51.2%) compared to males (48.8%). The majority of participants were aged 40–49 years (32.3%), with most having attained at least primary or secondary education.

**TABLE 1:** Socio-demographic Characteristics (n = 127)

| Variable | Category | Frequency | Percentage (%) |
| --- | --- | --- | --- |
| Sex | Male | 62 | 48.8 |
|  | Female | 65 | 51.2 |
| Age Group | 15–29 | 25 | 19.7 |
|  | 30–39 | 34 | 26.8 |
|  | 40–49 | 41 | 32.3 |
|  | ≥50 | 27 | 21.2 |
| Education Level | No formal | 18 | 14.2 |
|  | Primary | 40 | 31.5 |
|  | Secondary | 35 | 27.6 |
|  | Tertiary | 34 | 26.8 |
| Occupation | Unemployed | 30 | 23.6 |
|  | Self-employed | 55 | 43.3 |
|  | Formal employment | 42 | 33.1 |

**Table 2** shows the distribution of patient delay and related psychosocial and access factors. A substantial proportion of participants (59.8%) delayed seeking care for more than two months. High levels of perceived stigma (96.9%) and fear (66.9%) were reported. Distance to health facilities varied, with nearly one-third of participants living more than 10 km away.

**TABLE 2:**
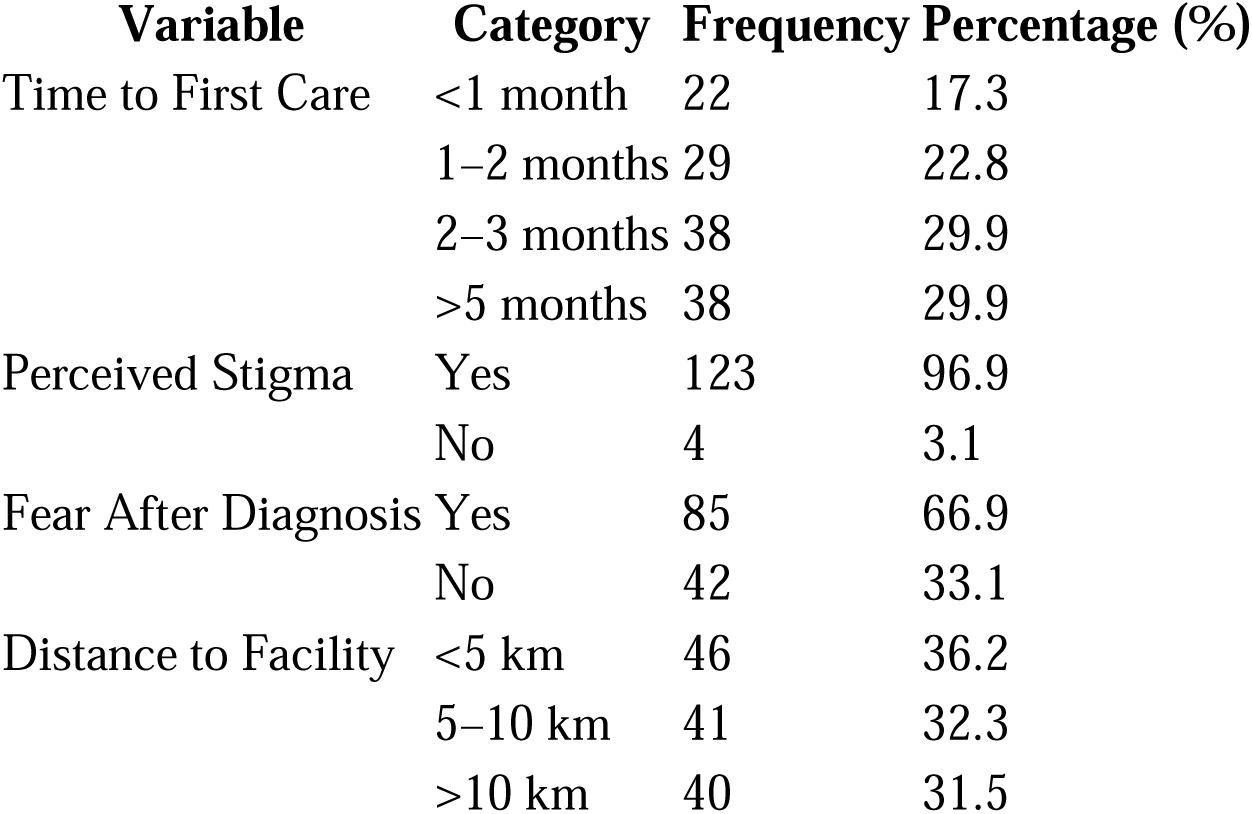
Patient Delay and Related Factors.

| Variable | Category | Frequency | Percentage (%) |
| --- | --- | --- | --- |
| Time to First Care | <1 month | 22 | 17.3 |
|  | 1–2 months | 29 | 22.8 |
|  | 2–3 months | 38 | 29.9 |
|  | >5 months | 38 | 29.9 |
| Perceived Stigma | Yes | 123 | 96.9 |
|  | No | 4 | 3.1 |
| Fear After Diagnosis | Yes | 85 | 66.9 |
|  | No | 42 | 33.1 |
| Distance to Facility | <5 km | 46 | 36.2 |
|  | 5–10 km | 41 | 32.3 |
|  | >10 km | 40 | 31.5 |

**Table 3** summarizes the bivariate associations between selected variables and delay in seeking care. Age group, distance to health facility, and education level showed statistically significant associations with delay (p < 0.05), while sex was not significantly associated.

**TABLE 3:** Bivariate Analysis of Factors Associated with Delay.

| Variable | Category | Delayed (%) | Not Delayed (%) | p-value |
| --- | --- | --- | --- | --- |
| Sex | Male | 60.0 | 40.0 | 0.210 |
|  | Female | 63.1 | 36.9 |  |
| Age Group | 15–29 | 52.0 | 48.0 | 0.032 |
|  | 30–39 | 58.8 | 41.2 |  |
|  | 40–49 | 70.7 | 29.3 |  |
| Distance | <5 km | 50.0 | 50.0 | 0.015 |
|  | >5 km | 68.0 | 32.0 |  |
| Education | Primary or less | 65.5 | 34.5 | 0.041 |
|  | Secondary+ | 55.2 | 44.8 |  |

**Table 4** presents the multivariable logistic regression analysis of factors associated with delay. Although some variables showed increased odds of delay, none were statistically significant (p > 0.05). This indicates that no independent predictors of delay were identified after adjusting for confounding variables.

**TABLE 4:** Multivariable Logistic Regression Analysis.

| Variable | Category | Adjusted OR | 95% CI | p-value |
| --- | --- | --- | --- | --- |
| Sex | Female vs Male | 1.21 | 0.65–2.25 | 0.540 |
| Age Group | 40–49 vs others | 1.48 | 0.78–2.81 | 0.210 |
| Distance | >5 km vs <5 km | 1.72 | 0.89–3.31 | 0.103 |
| Education | Low vs High | 1.36 | 0.72–2.56 | 0.330 |

Studies have shown that TB burden is disproportionately high among adults in their productive years, which contributes to sustained transmission within communities and significant socioeconomic consequences (World Health Organization, 2023; Furin et al., 2019). In high-burden settings, similar age distributions have been reported, reinforcing the importance of targeting TB control interventions toward working-age populations.

According to the World Health Organization, delays in seeking care contribute significantly to ongoing transmission and increased morbidity (WHO, 2023). Factors such as low awareness of TB symptoms, stigma, and reliance on self-medication have been widely documented as contributors to delayed care-seeking (Storla et al., 2008; Sreeramareddy et al., 2014). Prolonged patient delay increases the risk of disease progression and community transmission, highlighting the need for early detection strategies.

Fear of diagnosis and social consequences further compounds this delay, leading individuals to adopt concealment strategies or alternative care pathways (Courtwright & Turner, 2010; Craig et al., 2017). Additionally, geographic barriers such as distance to health facilities remain significant, particularly in resource-limited settings where access to healthcare services is uneven (Lönnroth et al., 2009).

These findings align with previous research demonstrating that TB-related stigma, fear, and structural barriers collectively contribute to diagnostic and treatment delays, ultimately sustaining transmission within communities.

Research indicates that patients often first seek care from pharmacies, traditional healers, or through self-medication before accessing formal healthcare services (Sreeramareddy et al., 2014; Hoa et al., 2013). Each step in this pathway introduces delays that contribute to late diagnosis and continued transmission.

This multi-step pathway underscores systemic inefficiencies and highlights the need for strengthening primary healthcare systems, improving referral mechanisms, and increasing community awareness to reduce delays in diagnosis and treatment initiation.

## DISCUSSION

Tuberculosis remains a significant public health challenge, with persistent delays in diagnosis and treatment driven by both patient-level and health system factors. A substantial proportion of participants in this study sought care more than two months after symptom onset, highlighting gaps in timely healthcare utilization and ongoing risks of transmission.

The findings indicate that tuberculosis disproportionately affects individuals in economically productive age groups, particularly those aged 40–49 years. This pattern is consistent with global epidemiological trends and has important implications for both disease transmission and socioeconomic burden. Delayed diagnosis within this population not only increases community spread but also contributes to loss of productivity and financial strain at household level.

Patient delay emerged as a major contributor to late diagnosis. This delay appears to be influenced by a combination of behavioral and structural factors, including limited awareness of tuberculosis symptoms, normalization of persistent cough, stigma, and barriers to accessing healthcare. These findings are consistent with previous studies conducted in similar high-burden settings, which highlight the role of patient behavior in delaying care-seeking.

Stigma and fear were prominent barriers affecting healthcare utilization. Fear of diagnosis, social exclusion, and discrimination may discourage individuals from seeking care early, leading to concealment of symptoms and prolonged delays. These findings reinforce the importance of addressing social determinants of health in tuberculosis control strategies.

Geographical access also played a role in influencing delay. Participants residing farther from health facilities were more likely to delay seeking care, reflecting ongoing inequities in access to health services. This underscores the need for decentralization of tuberculosis services and improved accessibility at community level.

The care-seeking pathways observed in this study were often complex and non-linear, with many participants initially engaging in self-medication or consulting informal providers before accessing formal healthcare services. Such fragmented pathways contribute to missed opportunities for early diagnosis and highlight inefficiencies within the health system. Strengthening referral systems and integrating informal providers into tuberculosis control efforts may help reduce these delays.

Although several factors were associated with delay at the bivariate level, no independent predictors were identified after adjustment. This suggests that delays in tuberculosis diagnosis are influenced by multiple interrelated factors rather than a single determinant, emphasizing the need for comprehensive and multi-sectoral interventions.

### Age Distribution and Its Implications (Objective 1: Patient Characteristics and Risk Profile)

The predominance of participants within the 40–49-year age group highlights that tuberculosis disproportionately affects individuals in their economically productive years. This finding aligns with global evidence indicating that TB primarily impacts adults who contribute significantly to household income and national productivity (World Health Organization, 2023; Furin et al., 2019).

From a public health perspective, this age distribution has important implications for both transmission dynamics and socioeconomic burden. Adults in this age group are more likely to engage in social and occupational interactions that facilitate disease spread, while also facing competing responsibilities that may delay healthcare seeking. This reinforces the need for targeted TB screening and health education strategies focusing on working-age populations.

### Patient Delay and Its Determinants (Objective 2: Health-Seeking Timeliness)

The study identified substantial patient delay, with many individuals seeking care more than two months after symptom onset. This delay is consistent with findings from systematic reviews showing that prolonged patient delay remains a major barrier to TB control in high-burden settings (Storla et al., 2008; Sreeramareddy et al., 2014).

Patient delay is often driven by a combination of low awareness of TB symptoms, normalization of chronic cough, reliance on self-care, and socioeconomic constraints. In addition, stigma and fear of diagnosis further discourage timely healthcare seeking (Courtwright & Turner, 2010).

These findings suggest that despite ongoing TB control efforts, early detection remains insufficient, allowing continued transmission within communities and increasing the risk of severe disease outcomes.

### Barriers to Care (Objective 3: Health System and Socio-Cultural Barriers)

The high prevalence of stigma, fear of diagnosis, and geographical barriers underscores the multifaceted challenges affecting access to TB care. Stigma continues to be a significant social determinant of TB outcomes, influencing individuals’ willingness to disclose symptoms and seek formal medical services (Craig et al., 2017).

Fear of social isolation, discrimination, and loss of employment often leads to concealment of symptoms, which delays diagnosis and treatment initiation. Additionally, distance to health facilities remains a structural barrier, particularly in resource-limited settings, where healthcare access is unevenly distributed (Lönnroth et al., 2009).

These findings highlight that TB control requires not only clinical interventions but also strong community engagement and stigma reduction strategies. Without addressing these barriers, delays in diagnosis and treatment will persist, undermining TB control efforts.

### Care-Seeking Pathways and Health System Inefficiencies (Objective 4: Patient Pathways to Care)

The observed care-seeking pathways reveal multiple points of delay, including self-medication, visits to informal providers, and eventual presentation to formal healthcare facilities. This fragmented pathway is widely documented in TB literature and reflects systemic inefficiencies in healthcare delivery (Hoa et al., 2013).

Patients often first seek care from pharmacies, traditional healers, or through informal channels before accessing formal diagnostic services. Each step introduces additional delays, contributing to disease progression and continued transmission (Sreeramareddy et al., 2014).

These findings emphasize the need for strengthening primary healthcare systems, improving referral systems, and integrating informal providers into TB detection and referral networks. Early detection strategies at community level could significantly reduce diagnostic delays and improve treatment outcomes.

### Limitations

This study has several limitations. First, recall bias may have affected the accuracy of reported symptom onset, although efforts were made to validate timelines using clinical records. Second, the study was conducted in a single tertiary facility, which may limit generalizability to other settings. Third, the retrospective design may be subject to incomplete records and missing data. Finally, potential selection bias may have occurred, as only patients who accessed care at KNH were included.

## RECOMMENDATIONS

Efforts to reduce tuberculosis delays should focus on strengthening early case detection, particularly among working-age populations. Community-based awareness programs are needed to improve recognition of TB symptoms and promote timely healthcare seeking. Addressing stigma through community engagement and psychosocial support is essential to reduce delays associated with fear and social barriers. Improving access to services through decentralization and strengthening referral systems, including engagement with informal providers, may further reduce diagnostic delays and improve treatment outcomes.

This study demonstrates that tuberculosis remains a significant public health challenge characterized by delays in diagnosis, persistent stigma, and fragmented care-seeking pathways. The predominance of cases among individuals in the 40–49-year age group underscores the continued impact of TB on economically productive populations, amplifying both health and socioeconomic consequences.

Patient delay was found to be a major contributor to late diagnosis, driven by low awareness, stigma, and reliance on informal care pathways. Barriers such as fear, stigma, and geographical inaccessibility further exacerbate these delays, while multiple care-seeking steps highlight inefficiencies within the health system.

Addressing these challenges requires a comprehensive, multi-sectoral approach that includes strengthening primary healthcare systems, enhancing community awareness, reducing stigma, and improving care integration. Such interventions are essential to achieving early diagnosis, reducing transmission, and ultimately advancing TB control and elimination efforts.

**Figure 1:**
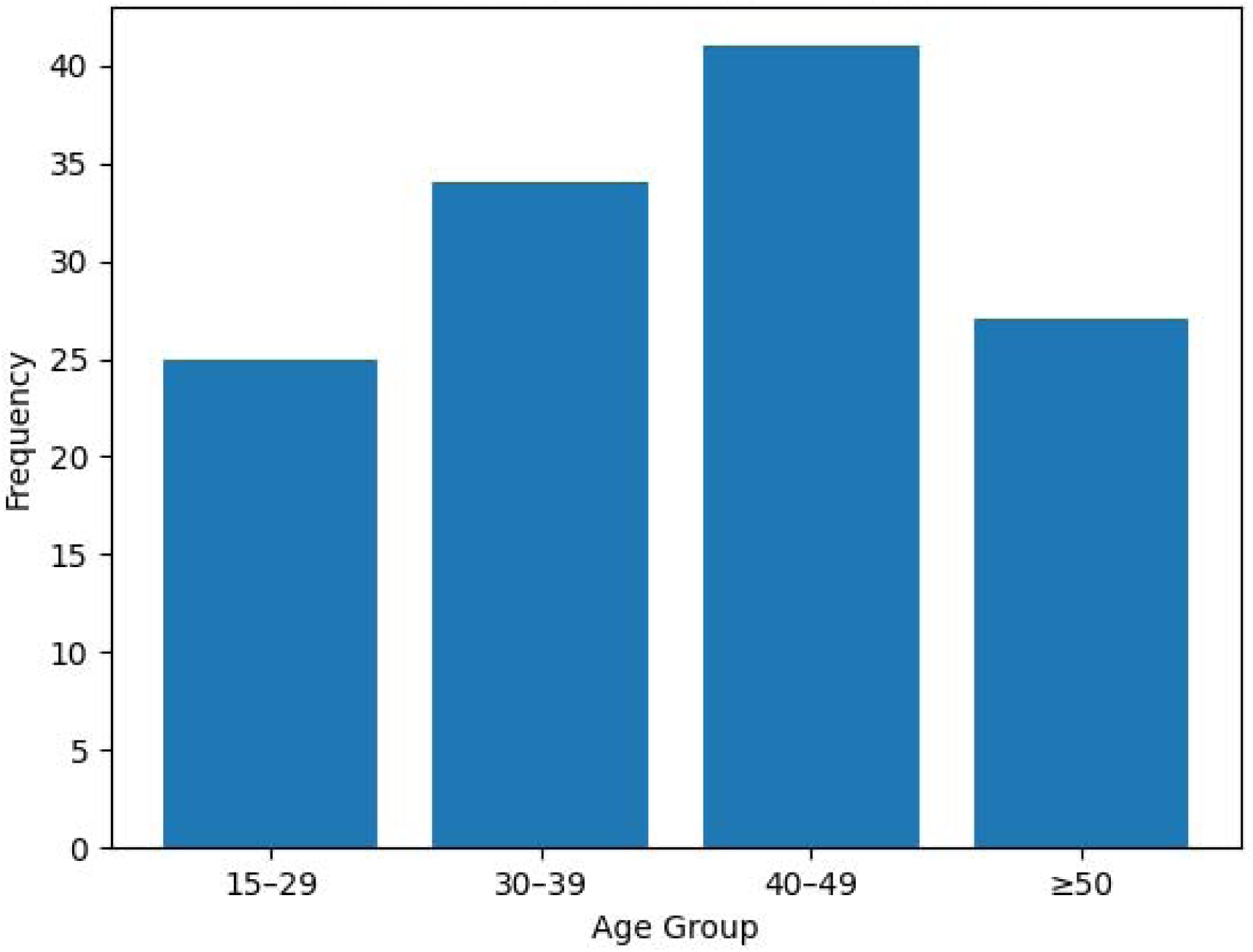
Age distribution of study participants (n = 127). *Figure 1* Shows highest concentration in 40–49 age group, indicating TB burden among middle-aged adults. The age distribution indicates that the highest proportion of participants were in the 40–49-year age group. This pattern is consistent with global tuberculosis epidemiology, where TB predominantly affects economically productive age groups. Individuals in this age range are often at increased risk due to cumulative exposure, occupational risks, and potential immunological compromise.

**Figure 2:**
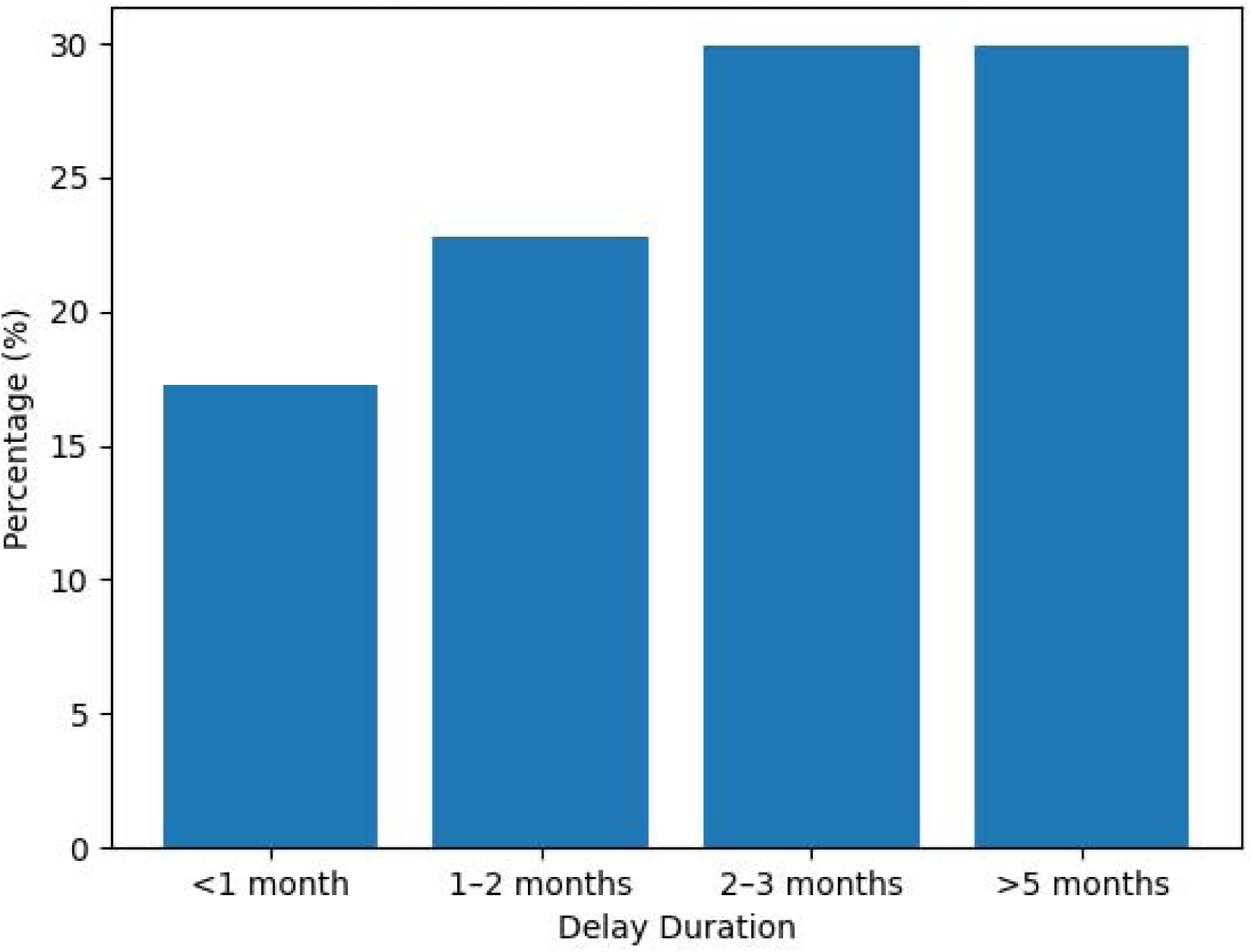
Distribution of patient delay in seeking care. *Figure 2 shows* Highlights prolonged delays, with many patients seeking care after more than two months. The findings reveal that a substantial proportion of participants experienced prolonged patient delay, with many seeking care after more than two months of symptom onset. This delay is consistent with evidence indicating that patient delay remains a major challenge in tuberculosis control, particularly in low- and middle-income countries.

**Figure 3:**
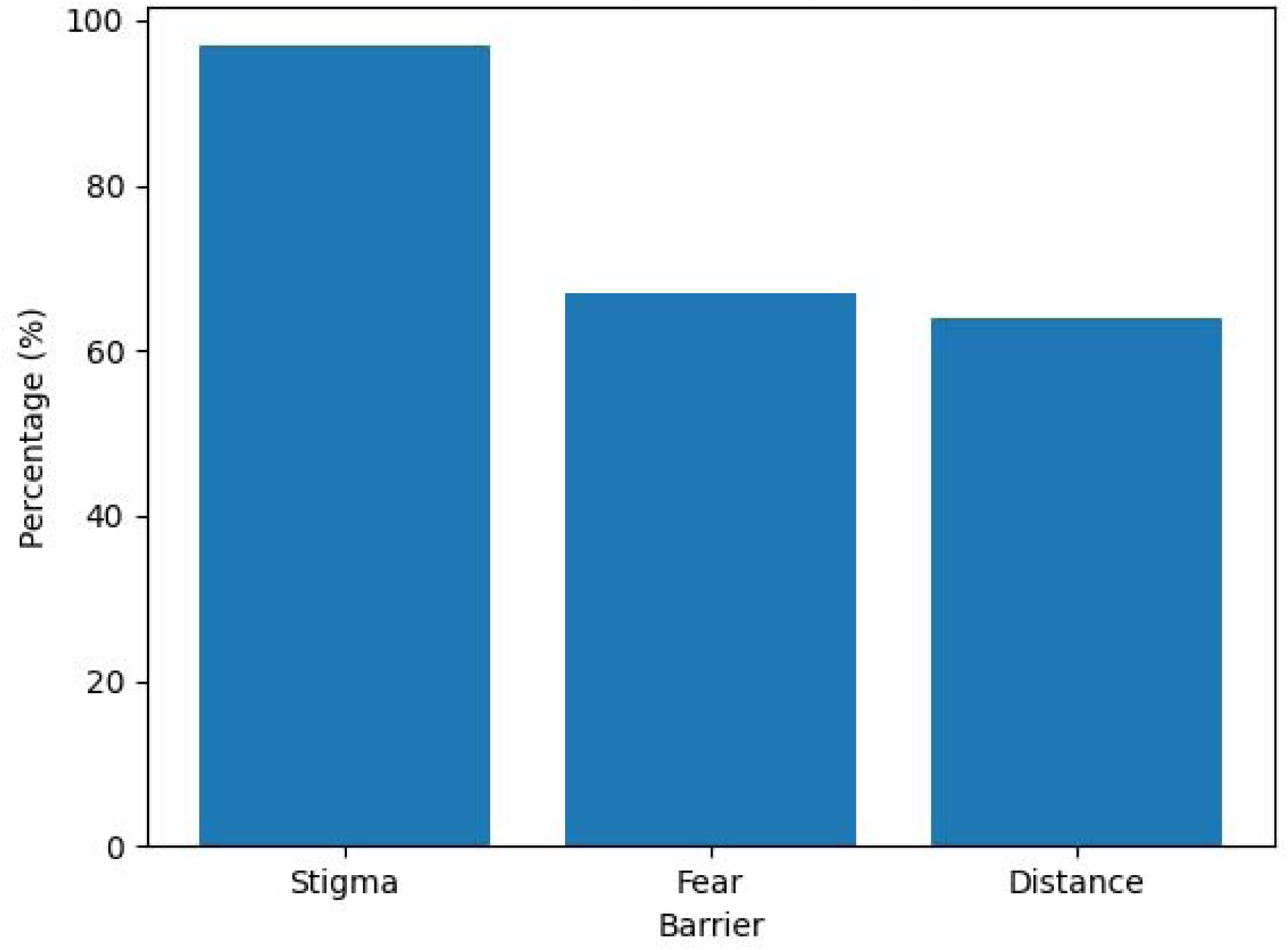
Reported barriers to timely care-seeking. *Figure 3*: Shows Stigma and fear are dominant barriers, followed by distance to health facilities. The high prevalence of perceived stigma among participants reflects a critical barrier to timely TB diagnosis and treatment. Stigma has been consistently identified as a major determinant of delayed care-seeking behavior, as it influences patients’ willingness to disclose symptoms and seek formal medical care.

**Figure 4:**
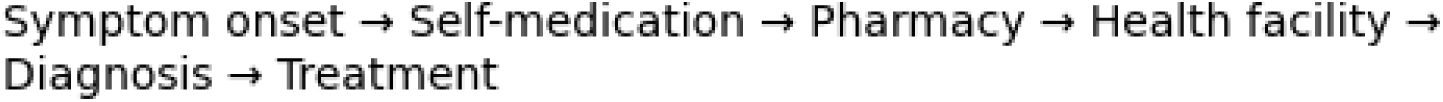
Care-seeking pathway prior to TB diagnosis. *Figures 4* Illustrates multiple steps before diagnosis, showing potential delay points. The care-seeking pathway illustrates multiple steps between symptom onset and treatment initiation, including self-medication and visits to informal providers before reaching formal health facilities. This pattern is widely documented in TB epidemiology and reflects fragmented healthcare-seeking behavior.

## Data Availability

All data produced in the present study are available upon reasonable request to the corresponding author.
This option is preferred because:
Patient-level TB data may involve confidential hospital records.
It complies with research ethics and data protection requirements.
Journals and conferences commonly accept this wording.
If the form requires a slightly more detailed version, you can use:
The datasets generated and/or analyzed during the current study are available from the corresponding author upon reasonable request, subject to ethical and institutional approval from Kenyatta National Hospital.

https://osf.io/7kq3p/

---

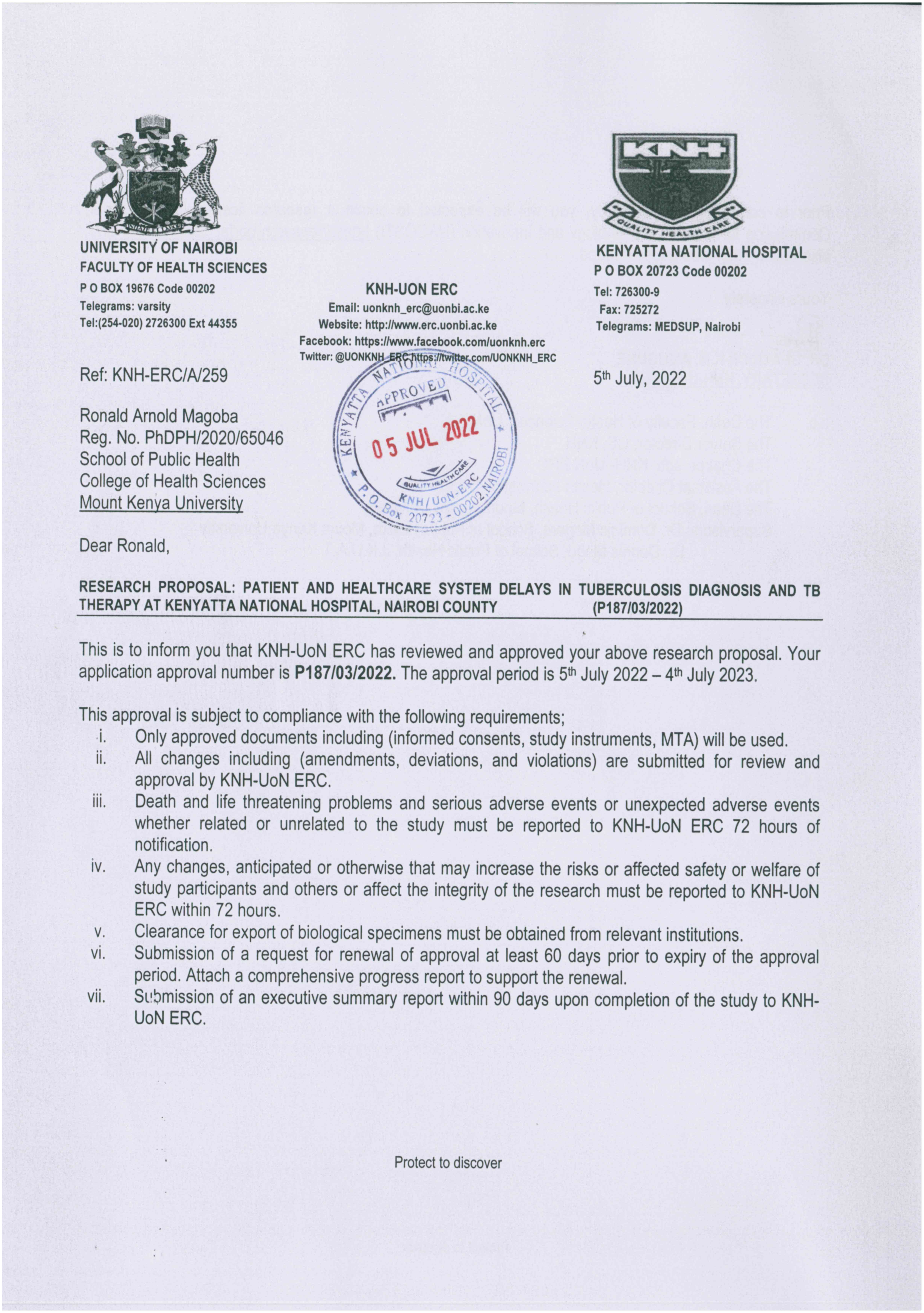
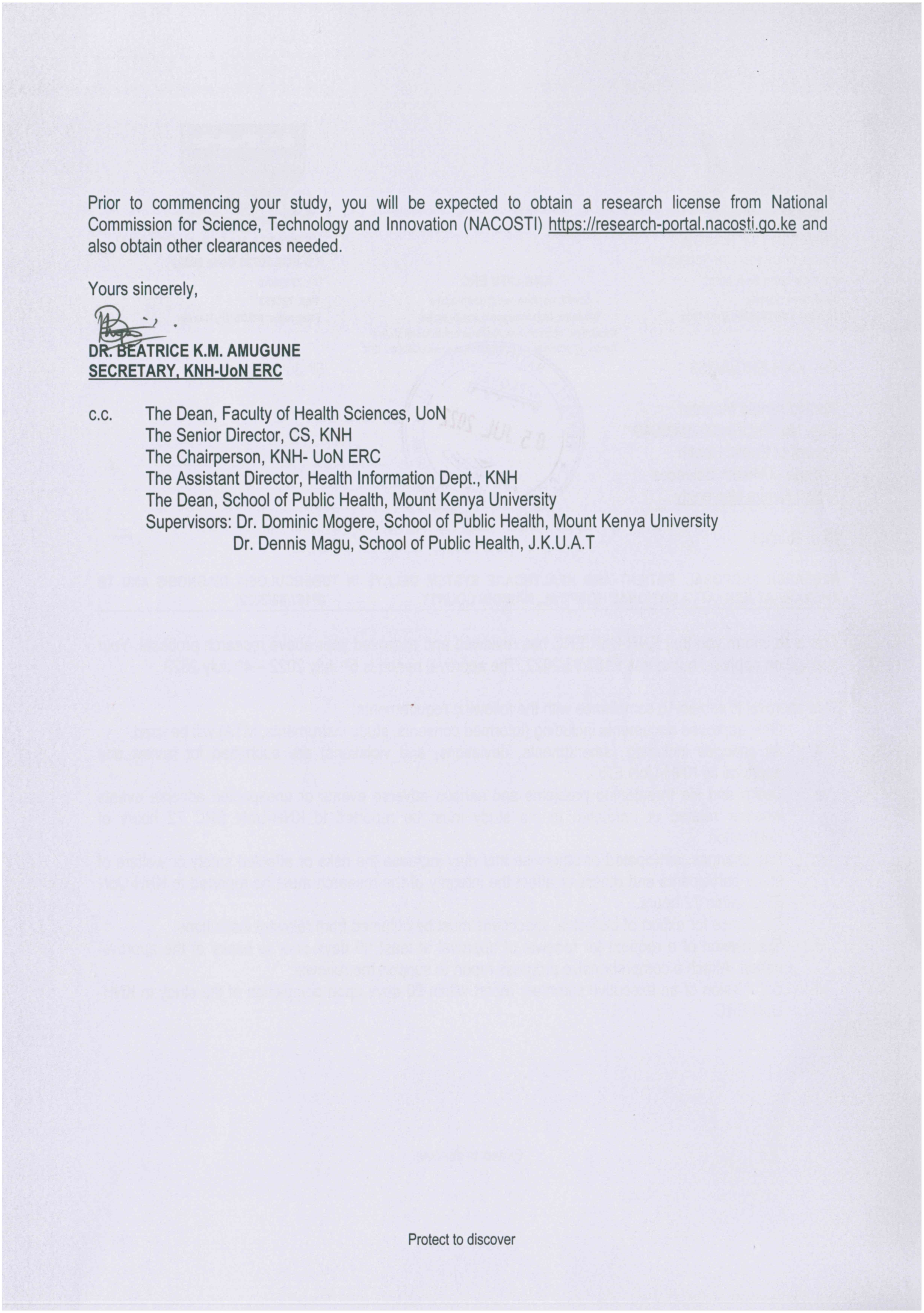

## REFERENCES

1. World Health Organization. Global tuberculosis report 2023. Geneva: WHO; 2023.

2. Storla DG, Yimer S, Bjune GA. A systematic review of delay in tuberculosis care. BMC Public Health. 2008; 8:15.

3. Sreeramareddy CT, Qin ZZ, Satyanarayana S, et al. Delays in diagnosis and treatment of tuberculosis. Int J Tuberc Lung Dis. 2014;18(3):255–266.

4. Courtwright A, Turner AN. Tuberculosis and stigmatization. Public Health Rep. 2010;125(Suppl 4):34–42.

5. Craig GM, Daftary A, Engel N, et al. Tuberculosis stigma. Int J Infect Dis. 2017; 56:150–159.

6. Lönnroth K, Castro KG, Chakaya JM, et al. Tuberculosis control and elimination. Lancet. 2009;375(9728):1430–1441.

7. Hoa NP, Chuc NT, Thorson A. Knowledge and practices about TB. BMC Public Health. 2013; 13:14.

8. Hoa NP, Chuc NT, Thorson A. Knowledge, attitudes, and practices about tuberculosis among rural communities in Vietnam. BMC Public Health. 2013; 13:14.

